# Variability of Individual Infectiousness Derived from Aggregate Statistics of COVID-19

**DOI:** 10.1101/2021.01.15.21249870

**Authors:** Julius B. Kirkegaard, Kim Sneppen

## Abstract

The quantification of spreading heterogeneity in the COVID-19 epidemic is crucial as it affects the choice of efficient mitigating strategies irrespective of whether its origin is biological or social. We present a method to deduce temporal and individual variations in the basic reproduction number *R* directly from epidemic trajectories at a community level. Using epidemic data from the 98 districts in Denmark we estimate an overdispersion factor *k* for COVID-19 to be about 0.11 (95% confidence interval 0.08 – 0.18), implying that 10 % of the infected cause between 70 % to 87 % of all infections.

---

In controlling epidemics, a deep understanding of the dynamics that underlie the spread of a disease is critical for choosing which interventions are most efficient to mitigate its continued spread. Epidemiological models of disease spreading [1, 2] depend on parameters that capture effects both of the pathogen–host biology and the behaviour of the population in which the disease propagates. Population-level data allow the estimation of the *basic reproduction number R*, denoting the average number of people an infectious individual will transmit the disease to. Hidden in the average value of *R* are both temporal variations and variations between infectious individuals [3]. Variations in time stem both from the fact that social behaviour can change during an epidemic due to e.g. interventions being put in place, and because as the epidemic progresses the susceptible fraction of the population decreases. Variations from person to person can results both from biological differences or social behaviour.

A popular tale for some diseases is the 20/80-rule stating that 20 % of infectious individuals are responsible for 80 % of all infections. This was for example seen in recent epidemics such as the 2003 Asia outbreak of SARS [3] and the 2014 Africa outbreak of Ebola [4]. Numerous of studies of COVID-19 suggest even more extreme statistics for this disease [5–9]. These effects have collectively becomes known as *superspreading*, and while it is simple to define theoretically, measuring them typically requires data at the level of individuals. Viral genome sequences can be used to inform the analysis [10, 11], and when contact tracing data is available [12–14] the analysis may be performed directly. More indirectly, the number of imported versus local cases has also been shown to inform the dispersion [15].

Here we derive a Bayesian model that use readily available time-resolved data at the population level to estimate infection heterogeneity. The typically large number of cases in country level data results in aggregate data that mask variations, however. But for the on-going COVID-19 epidemic many countries report their case statistics at a regional level. Here we show how this can be exploited to estimate dispersion parameters. Our method is particularly strong when the sizes of the regions vary significantly. The data from large regions constrain the average behaviour of *R*, whereas variations in the data from the smaller regions constrain variations in *R*.

We apply our approach on data for Denmark, which has a number of features that permit the analysis: Denmark, with a population of 5.8 million, makes available daily data for all of its 98 municipalities, which all coordinate their testing identically. As shown in Fig. 1(a – f), the number of cases in these municipalties vary significantly. In the capital region of Copenhagen, daily cases number in the hundreds, whereas in more rural Vesterhimmerlands the daily rate is less than ten. Finally, the population of Denmark is fairly uniform and thus slow, temporal variations in *R* can be assumed to affect all regions.

**FIG. 1.**
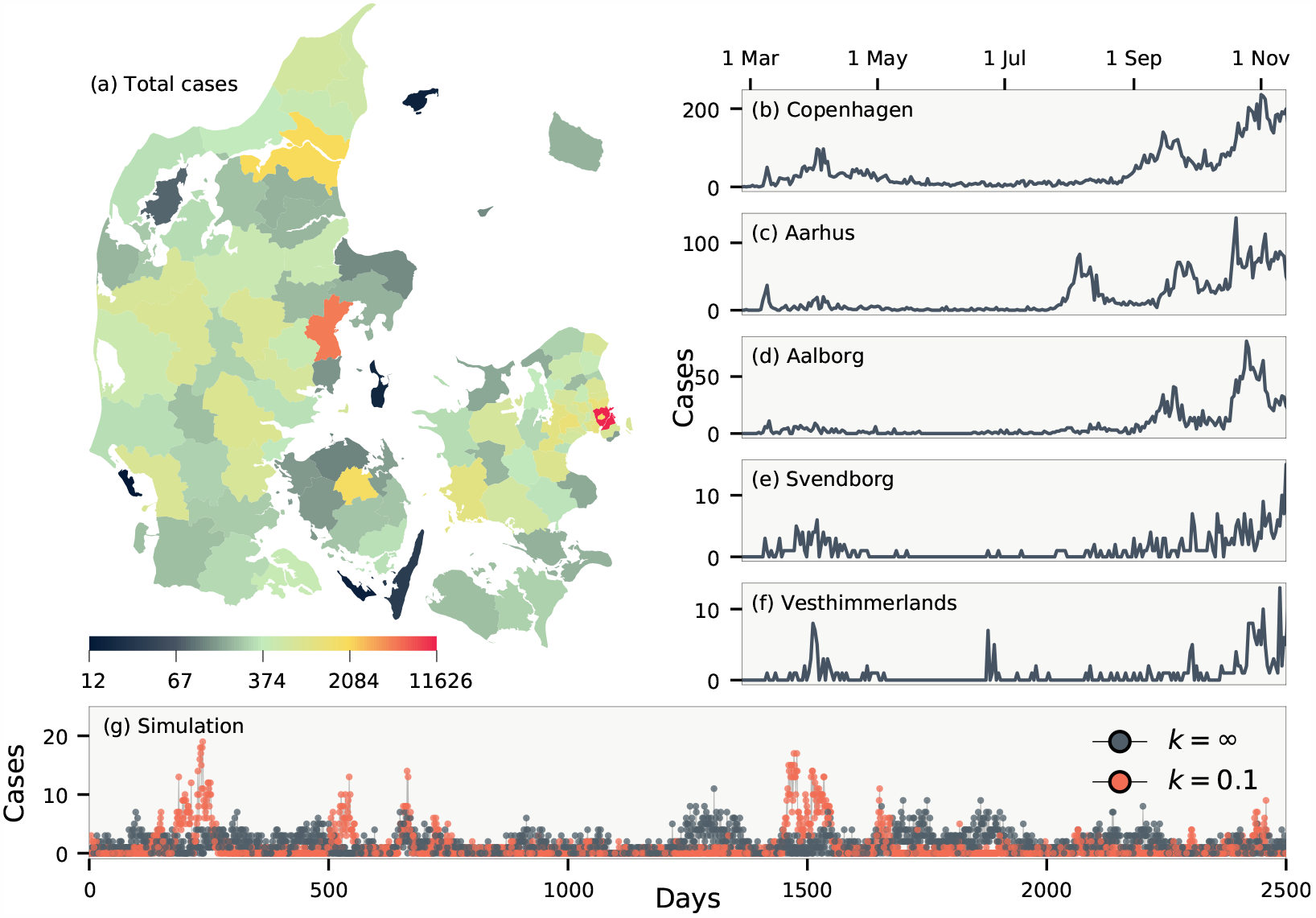
Daily cases of COVID-19 in Denmark. (a) The total number of cases in each of Denmark’s 98 municipalities. (b-f) Daily number of cases in five municipalities. (g) Simulations of an epidemic with dispersion parameter *k* = ∞ and *k* = 0.1, respectively. Both simulations use *R* = 0.9 and a crossing parameter chosen such that on average an infectious person enters every fifth day.

Following Lloyd-Smith et. al. [3], we assign each person an infectivity *ν* sampled from a gamma distribution Γ(*R, k*) with mean *R* and dispersion parameter *k*. Small *k* correspond to a disease driven mainly by superspreading as illustrated in Fig. 2(a). Accounting for subsequent independent stochastic infections, the offspring distribution is negative binomial NB(*R, k*) [3]. The parameters in this distribution can be deduced if contact tracing data is available. Using only aggregate data, however, we need to instead build a probabilistic augmentation of the missing contact information.

**FIG. 2.**
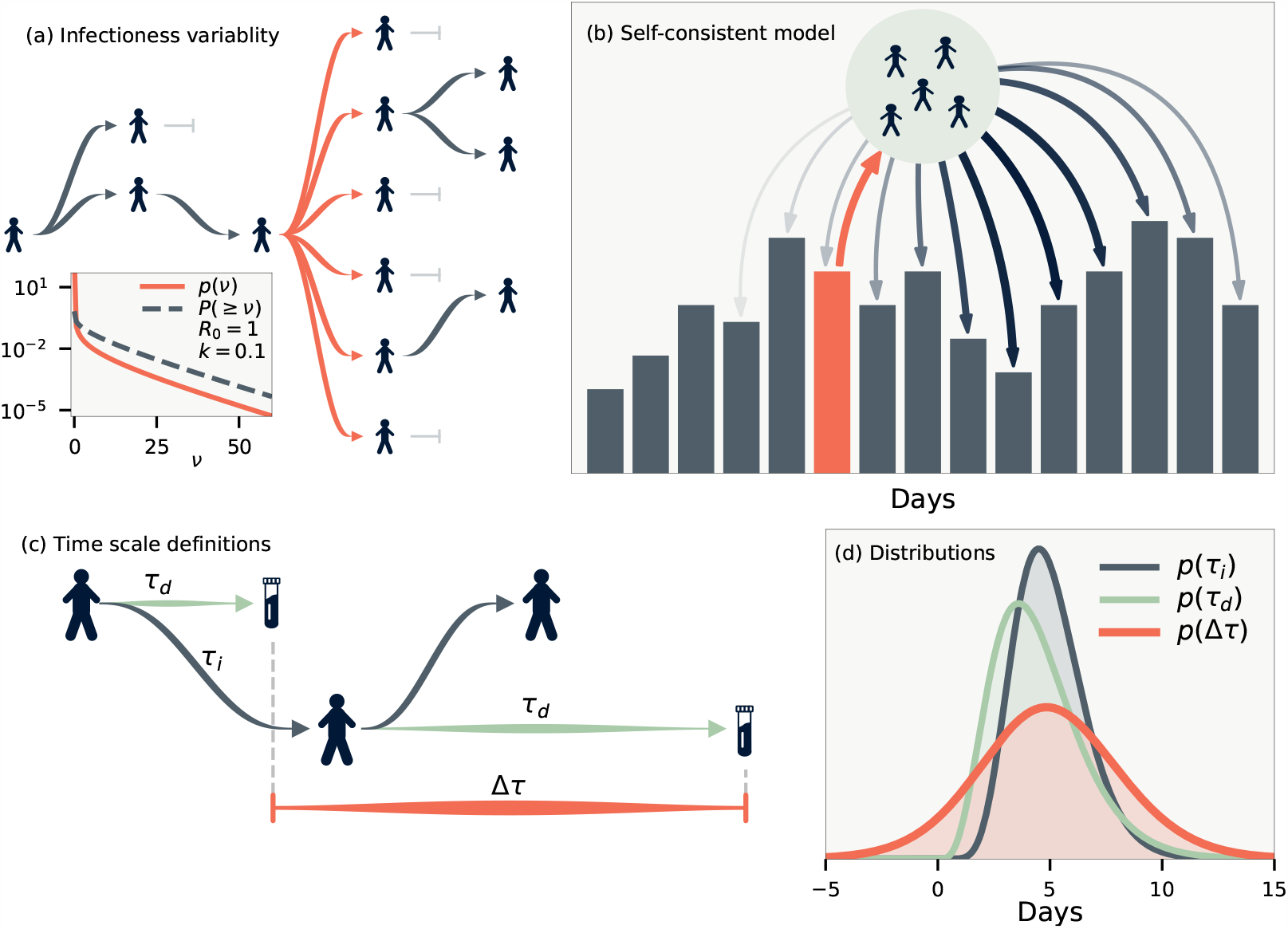
Model definitions. (a) Illustration of a heterogeneous infection pattern (superspreading). Inset shows the probability density function and (one minus) the cumulative probability for the gamma distribution Γ(*R* = 1.0, *k* = 0.1). (b) Likelihood model. The infected individuals who were discovered on some day (orange) will themselves infect a number of people. These are in turn detected on other days according to the distribution *p*(Δ*τ*). (c) Time scale definitions. *τ*_*d*_ denotes the duration from being infected to being discovered, and *τi* the duration between infections (generation time). Finally, Δ*τ* is the time between the discoveries of infector-infectee pairs. (d) The maximum likelihood of the distributions we employ for *τ*_*d*_ and *τ*_*i*_, and the distribution thus implied for Δ*τ* . *p*(Δ*τ*) has support below zero as it is possible that the infector is discovered after the infectee.

In aggregate data, the time between the discovery of an infector-infectee pair Δ*τ* is stochastic. This distribution can be calculated if the distribution of infection-to-infection *p*(*τ*_*i*_) (generation time) and infection-to-discovery *p*(*τ*_*d*_) are known. As illustrated in Fig. 2(c), the time between discoveries obey the random variable relation Δ*τ* = −*τ*_*d*_ + *τ*_*i*_ + *τ*_*d*_, where the first *τ*_*d*_ refers to the random time of discovery for the infector and the latter *τ*_*d*_ the time of discovery for the ∼ ± ± ∼ ± ± infectee. These do not affect the mean value of Δ*τ* but do increase its variance. The resulting distribution is shown in Fig. 2(d) using estimates from the literature of *p*(*τ*_*i*_) ∼ Γ(5.0 0.75, 10 1.5) and *p*(*τ*_*d*_) ∼ Γ(4.5 0.75, 5.0 1.0) [16, 17]. With these distributions in place it is straightforward to simulate an epidemic if *R*(*t*) and *k* are known. Fig. 1(g) shows two such examples for *k* = ∼ and *k* = 0.1. For *k* = 0.1 there will be superspreading, but because of the *∼* distribution of Δ *τ* these will be distributed over a number of days, making statistical analysis crucial for its discovery.

To tackle the inverse problem of the simulation we define a self-consistent model of the data. For simplicity let us first assume that all infectious individuals are found and postpone the discussion of under-reporting. Fig. 2(b) illustrates our approach: we define the likelihood of the data by calculating the probability of the observed time-series for each municipality. In practice, this can only be calculated in a reasonable amount of time because of a few key features of the negative-binomial distribution. These are derived in the SI, but can be summarised as follows: If the offspring distribution from a single individual is the negative binomial NB(*R, k*), then the offspring distribution from *M* people, where each individual is found on one specific day with probability *p* is exactly NB(*p MR, Mk*). The total likelihood of a single day is then found by convolving these distributions using (*p*Δ*τ*) for the daily probability of discovery. The precise formulae are presented in the SI.

To complete our model, we need to adjust for correlations that are present in the data as shown in Fig. 3. Naturally, a municipality with a large population will have a larger number of cases per day than a municipality with a small population. This is because there will be more imported cases in large regions (there may also be variations in *R* between cities and rural areas [18], but this is a second-order effect that we ignore). As most imported cases will come from other municipalities, we ignore effects of international travel. In fact, daily cases per population of the municipalities will be strongly correlated as a function of time as demonstrated in Fig. 3(a), reflecting the fact that Denmark is a small country with overall homogeneous development of the disease. To account for coupling between communities we introduce a crossing parameter *c* that corrects for the fraction of infections that occur across municipality borders. For the number of infectious individuals in municipality *m* we thus use 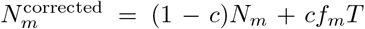, where *N*_*m*_ is the uncorrected number of infectious individuals in municipality *m, f*_*m*_ is the population fraction of municipality *m*, and *T* is the total number of infectious individuals across all municipalities. With this simple formula it is ensured that municipalities will, on average, have a number of infections that is proportional to the population of the municipality.

**FIG. 3.**
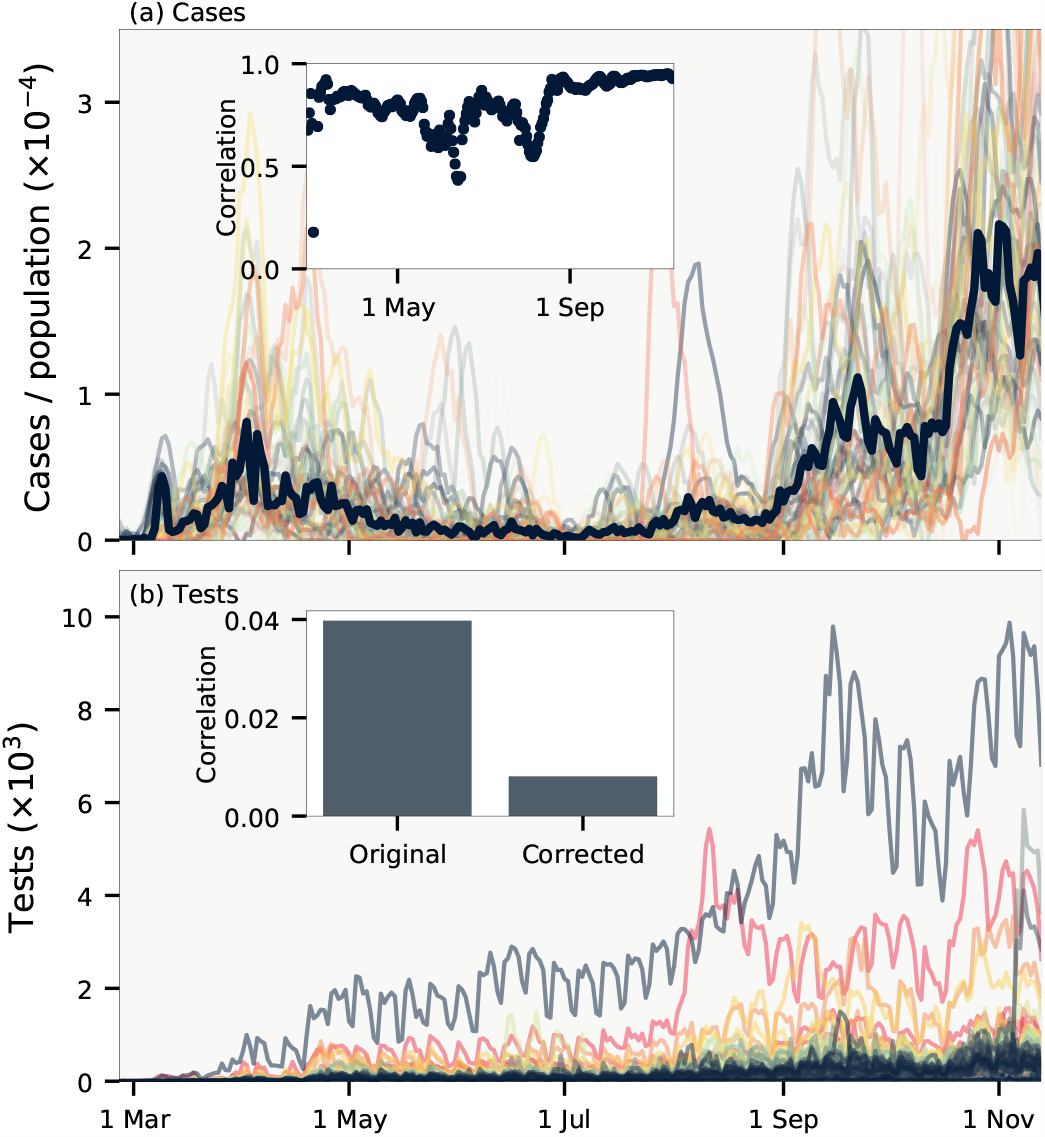
Data correlations. (a) Cases per population of each municipality smoothed over one week. Thick line shows cases for all of Denmark. Inset shows the cross correlation between municipalities as a function of time. (b) Daily test frequency in each municipality. Inset shows the correlation of deviations of daily cases from a weekly running mean both with and without linear correction for the number of tests.

Fig. 3(b) shows that the testing frequency in each municipality is highly irregular, with e.g. fewer tests being done on weekends. Our method to detect variations in reproduction number depends on the deviations in cases in each municipality to be uncorrelated. The inset of Fig. 3(b) shows that there is a small correlation present. This is natural, since the number of tests is correlated across municipalities. We correct for this effect by scaling with the number of tests. This is incorporated into our model by re-scaling the distribution of discovery *p*(*τ*_*d*_) in proportion to the daily number of tests (see SI for details).

Finally, we employ Hamiltonian Monte Carlo [19] to sample for *R*(*t*), *k* and *c* from the total likelihood function of all regions, aimed to reproduce the case counts at each day, given case counts on previous days. In particular, we run the NUTS algorithm [20] with gradients of the log likelihood calculated by automatic differentiation [21] on GPUs that allow for fast calculations of convolutions that make up our likelihood function (see SI). We restrict temporal variations of *R*(*t*) to be slow on the scale of weeks by parameterising the function using cubic Hermite splines. The Hamiltonian Monte Carlo chain is then run multiple times for sampled *p*(*τ*_*i*_) and *p*(*τ*_*d*_).

Our results are shown in Fig. 4. The sampling reveals an *R*(*t*) [Fig. 3(a)] that slightly deviates from estimates obtained by single approximations using e.g. the SIR model [1, 22]. This is because we calculate an *R*(*t*) that best explain the statistics of each municipality and not the sum of these. Further, we have a large uncertainty on our estimates because our *R*(*t*) models the reproduction number under uncertain values of *p*(*τ*_*i*_) and *p*(*τ*_*d*_) (see Ref. [23] for details on precise estimation of *R*(*t*) alone). In other words, we calculate the true value of *R* as defined by the average offspring count, and not as the value of *R* that best makes a single model fit the evolving infection statistics [24].

**FIG. 4.**
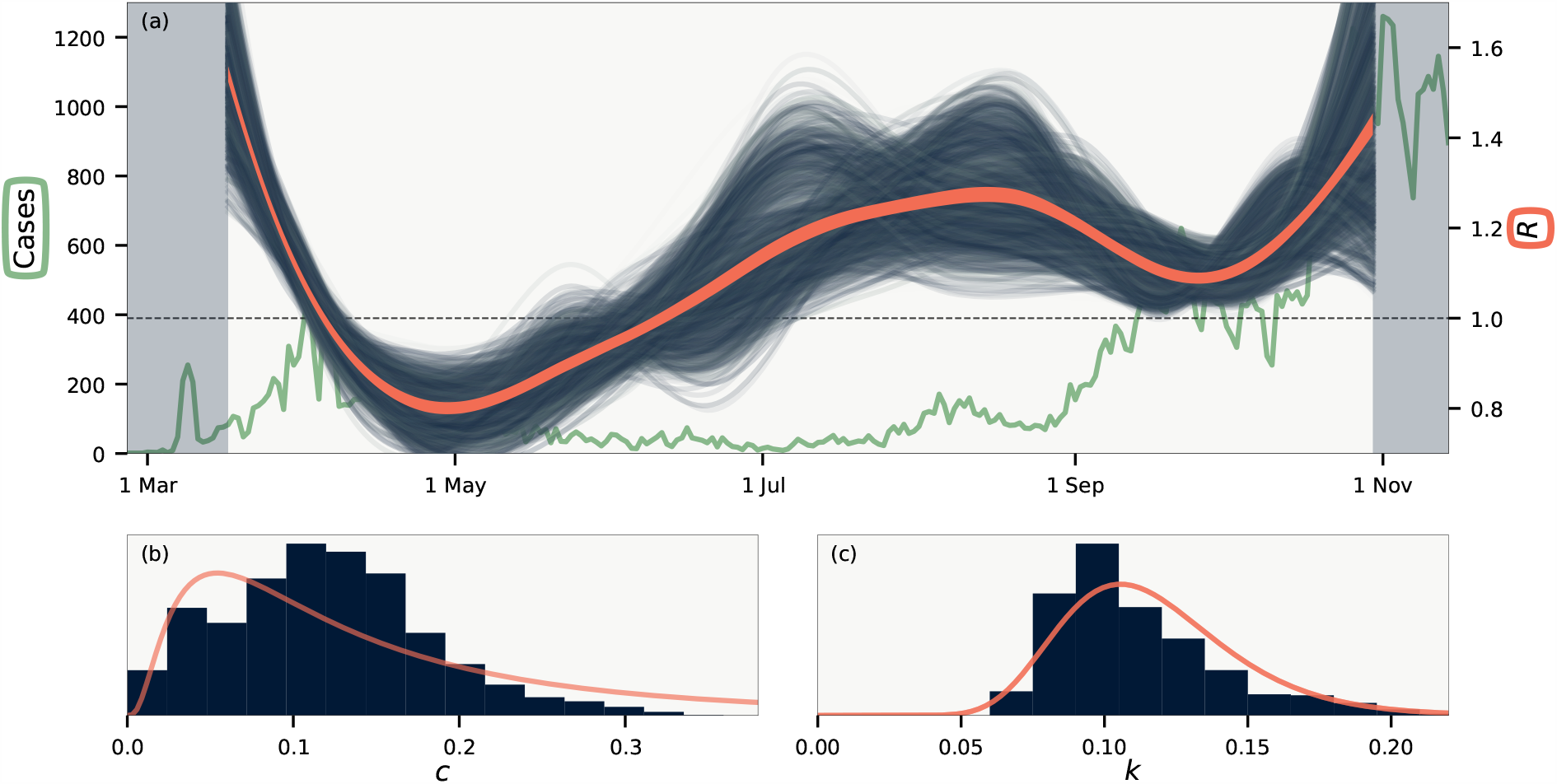
Results. (a) Temporal variations of the basic reproduction number *R*(*t*). Background line shows total number of daily cases. Blue lines are realisations from the MCMC sampling, while orange line indicates average of all samples. Shaded background shows sections of the data that are not included in the likelihood. (b) Histogram of the crossing parameter *c*. (c) Histogram of the dispersion parameter *k*. Curves in (b–c) are log-normal distributions with matching mean and variance.

Fig. 3(b) shows that we cannot constrain *c* more than to say that by far most infections happen within municipality borders, as is expected. In contrast, the degree of superspreading as defined by the value of *k* is fairly constrained as shown in Fig. 4(c). We find *k* in the range 0.08 – 0.18 (95% confidence interval), with mean *k* = 0.11, which compares well to estimated confidence intervals obtained from other methods [10, 12, 13, 15] but smaller than *k* ∼ 0.4 reported by Ref. [14]. For *R* = 1.4, for instance, our value range corresponds to an epidemic in which 10 % of the infected individuals are responsible for 70 % – 87% of all cases. In this case, the majority of infectious individuals will not infect anyone, in broad agreement with the fact that there are remarkably few transmissions within households [25, 26]. We note that the precise range of such statistics depends on the choice of probability distribution for infectiousness, for which we used the gamma distribution as has become standard [3]. If, for instance, the distribution instead were fat-tailed then other statistics would emerge [7, 27]. The value of the crossing parameter *c* only weakly affects *k*, which is instead affecting mainly by the mean value of *τ*_*i*_ and the widths of the distributions of *τ*_*i*_ and *τ*_*d*_. In particular, in our model we assume that infectious individuals spread the disease over time. If, in contrast, the spread from individuals is driven mainly single events then our distribution for *p*(Δ*τ*) is too wide. To study the effect of this, we ran our model with both *p*(*τ*_*i*_) and one of the two *p*(*τ*_*d*_) that make up *p*(Δ*τ*) constrained to a single day. This leads to a *k* that is about 40 % larger than the one estimated.

We have until now assumed that all infectious individuals were included in the data. This is of course not true. Focusing on estimating *k* we here consider the case where only a (time-independent) fraction *f <* 1 of all infectious are found. This leads our method to overestimate *k*. Most simply, if the incidence at each day is a factor 1*/f* larger than the measured data, fluctuations are amplified by 1*/f* and the true dispersion parameter *k* will be our measured *k* multiplied by *f* . Thus a value of *k* = 0.1 from Fig. 4c and an *f* ∼ 1*/*3 would correspond to a true *k* ∼ 0.03. It is however more realistic to assume that each detected case is independently found with probability *f* . Using simulated data where a fraction *f* ∼ ⅓ of cases are independently detected we find that a measured *k* of 0.1 correspond to a true underlying *k* that is between 0.05 and 0.085, depending on the simulation. If, on the other hand, there is large correlations between the reporting present in the data, our method may underestimate *k*.

These systematic uncertainties should be considered for our estimated value of *k*. The existence of large spreading events makes our model underestimate *k*, whereas uncorrelated under-reporting leads to overestimation. The effects will tend to cancel each other, but taken to the extreme could bring *k* to 0.04 – 0.28. We have furthermore tested our method on random subsets of all municipalities, and found that this did not have any significant impact on our estimates.

Traditionally one characterises an epidemic with only one number, *R*_0_, and even so there are remarkably few direct measurements of this average for known diseases. Here we ventured beyond such average measurements and proposed a new community level method to extract also variations in infectivity without having access to person sensitive data and contact tracing. Using our method we quantified the COVID-19 epidemic as one of the most extreme superspreader dominated diseases ever recorded [3]. This implies that it should be comparatively easy to mitigate with societal restrictions [8, 9].

## Data Availability

Data is publicly available.

## Acknowledgments

This project has received funding from the Novo Nordisk Foundation, under its Data Science Initiative, Grant Agreement NNF20OC0062047, and from the European Research Council (ERC) under the European Union’s Horizon 2020 Research and Innovation Programme, Grant Agreement No. 740704.

