## Supplemental Text for "Variability of Individual Infectiousness Derived from Aggregate Statistics of COVID-19"

Julius B. Kirkegaard and Kim Sneppen
*Niels Bohr Institute, University of Copenhagen, 2100 Copenhagen, Denmark*
(Dated: January 15, 2021)

### BASIC DISTRIBUTIONS

The offspring distribution from an individual with infectivity $\nu$ is Poissonian:

$$p(n) = \mathrm{Pois}(n;\, \nu) = \frac{\nu^n\, e^{-\nu}}{n!}. \tag{1}$$

When infectivty $\nu$ is distribution according to a Gamma distribution

$$p(\nu) = \Gamma(\nu;\, R, k) = \frac{\nu^{k-1}}{\Gamma(k)} \left(\frac{k}{R}\right)^k e^{-\frac{k\nu}{R}}, \tag{2}$$

the total offspring distribution becomes negative binomial

$$p(n) = \mathrm{NB}(n;\, R, k) = \frac{\Gamma(n+k)}{n!\,\Gamma(k)} \frac{k^k R^n}{(k+R)^{n+k}}. \tag{3}$$

### NEGATIVE BINOMIAL FORMULAS

The negative binomial has probability generating function

$$G_n(s; R, k) = \left[1 + \frac{R}{k}(1-s)\right]^{-k}. \tag{4}$$

The number of infections from $M$ infectious people will then have generating function

$$G_n^M(s) = \left[1 + \frac{R}{k}(1-s)\right]^{-Mk} = \left[1 + \frac{MR}{Mk}(1-s)\right]^{-Mk} = G_n(s; MR, Mk). \tag{5}$$

If a person is only discovered with probability $p$, corresponding to a Bernoulli random variable with generating function $G^B(s) = ps + (1-p)$, the generating function for the discovered offspring distribution becomes

$$G_n^D(s) = G_n\big(G^B(s)\big) = \left(1 + \frac{pR}{k}(1-s)\right)^{-k} = G_n(s; pR, k). \tag{6}$$

These formulas combined show that the discovered offspring distribution from $M$ people is $\mathrm{NB}(pMR, Mk)$.

### BASE LIKELIHOOD MODEL

We derive our likelihood model by calculating the probability to observe a given number of cases on a specific day given the previous days' case counts.

Define the variable $z(d_1, d_2)$ as the number people discovered on day $d_2$ whose infector was discovered on day $d_1$. Using the above results we have

$$z(d_1, d_2) \sim \mathrm{NB}(p(d_2 - d_1)\, c_1\, R, c_1\, k), \tag{7}$$

where $c_1$ is the number of cases on day $d_1$, and $p(\Delta\tau)$ is the distribution of the time between discoveries.

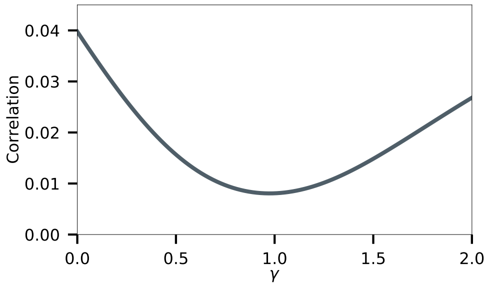


FIG. 1. Correlation of deviation in cases as a function of correction exponent $\gamma$. Best correction is found for $\gamma \approx 1$.

The number of infections $c_2$ on day $d_2$ is given by

$$c_2 = \sum_{d_1} z(d_1, d_2). \tag{8}$$

In other words: the number of infectious discovered on day $d_2$ is the sum of those cases from the surrounding days ($\{d_1\}$) that are discovered on day $d_2$. We make the assumption that these are independent, although this is not strictly true. In this case $c_2$ will be distributed as

$$c_2 \sim \underset{d_1}{\circledast} \mathrm{NB}(p\,(d_2 - d_1)\, c_1\, R, c_1\, k), \tag{9}$$

where $\circledast$ denotes convolution.

The total log likelihood is then found by summing over all regions and days:

$$\log \mathcal{L} = \sum_{\text{regions}} \sum_i \log\left( \underset{j}{\circledast} \mathrm{NB}(c_i\,;\, p\,(d_i - d_j)\, c_j\, R,\, c_j\, k) \right), \tag{10}$$

where $c_i$ is the number of cases on day $d_i$. We use PyTorch to evaluate this expression and its gradients on an Nvidia Geforce RTX 2080 Ti GPU.

### CORRECTION FOR NUMBER OF TESTS

To correct for the number of tests in the distribution of

$$\Delta\tau = d_2 - d_1 = \tau_2 + \iota_2 - \tau_1, \tag{11}$$

we rescale $p(\tau_d)$ by the number of tests. Denote by $T(d)$ the number of tests on day $d$, then we redefine

$$p(\tau_d, d) = \frac{p(\tau_d)\, T(d)}{\sum_\delta p(\tau_d + \delta)\, T(d + \delta)}. \tag{12}$$

This allows us to calculate an updated version of $p(\Delta\tau)$ which now must be written as

$$p(d_1, d_2) \tag{13}$$

as it explicitly depends on the days.

To verify that this is the right correction to make, we consider, as discussed in the main text, the correlation between regions of the deviations of daily cases from a running mean. In the main text it is shown that $c_i(d) - \langle c_i(d)\rangle$ has a larger correlation coefficient than $c_i(D)/T_i(d) - \langle c_i(d)/T_i(d)\rangle$. One would generally not expect the number of discovered cases to scale direct in proportion to the number tests, however. Thus we also considered the correlation of $c_i(d)/T_i(d)^\gamma - \langle c_i(d)/T_i(d)^\gamma\rangle$ As shown in Fig. 1, we find the smallest correlation for exactly $\gamma \approx 1$ when we compare to a running mean on the scale of weeks.

### TOTAL LIKELIHOOD

Using $c_i(d)$ for the number of cases in region $i$ on day $d$, our total log likelihood is calculated as follows:

1. Assign parameters $R(t)$, $k$ and $c$.
2. For each region $i$:
   (a) Calculate the effective number of cases taking crossing into account: $c_i^{\text{eff}} = (1-c)\,c_i + cf_iC(d)$, where $f_i$ is the population fraction and $C(d)$ the total number of cases across all regions on day $d$.
   (b) For each day $d$:
      i. Calculate $p(d', d)$ using Eq. (12)
      ii. Use $p(d', d)$ in Eq. (9) to calculate $p(c_i^{\text{eff}}(d))$
3. Take the logarithm of $p(c_i^{\text{eff}}(d))$ and sum over all days and regions.

### CHECKS & CAVEATS

To check our method we compared it to a simulation. In the simplest case, this simulation was a run as a $\tau$-leaping stochastic simulation taking steps of one day:

1. Assign $R(t)$, $k$, $p(\tau_i)$ and $p(\tau_d)$.
2. Initialise a list of current infectious people and their infectivity $\boldsymbol{\nu} = [\nu_1]$ and day on which they got infected $\boldsymbol{d} = [d_1]$
3. For each day $d$
   (a) Calculate number of people infected on this day as $n_d = \sum_i X_i$, where $X_i$ is the number of people that person $i$ infects on day $d$, sampled from $\text{Pois}(\nu_i \cdot p(\tau_i = (d - d_i)))$.
   (b) For each newly infected people, update $\boldsymbol{\nu}$ and $\boldsymbol{d}$ with new infectivities $\nu_j$ sampled from $\Gamma(R(d), k)$.
4. For each person, sample their day of discovery from $p(\tau_d)$.

We ran these simulations using 100 regions over 250 days, matching the data used in the main text. Using our likelihood model, we were consistently able to reproduce $R$ and $k$ to within a few percent of the simulations despite the assumptions used to derive our model. We also tested the effect of including imported cases using a crossing parameter. Finally, we subsampled our simulations using a binomial distribtion to simulate the effects of under-reporting.

To test the dependency of our results on specific municipalities we ran our model with only half of all municipalities sampled randomly. All test such test runs returned similar results. We further tested explicitely excluding the capital region (Copenhagen, Frederiksberg, Gentofte, Dragør, Vallensbæk, Gladsaxe, Herlev, Hvidovre, Rødovre, Brøndby, Glostrup, Høje, Taastrup Tarnby, Ishøj and Albertslund) which again did not make a significant difference.

Restricting our model to subsets of the full time interval naturally results in variation. Solving for the maximum likelihood of $R$ and $k$ we found variations not just for $R$, but also the dispersion parameter $k$, which (for fixed distributions of $\Delta\tau$) varied between $0.08 - 0.16$.

In closing we note that our analysis depends on the following assumptions:

- We assume the value for $R$, $k$, $c$ and distributions of $\Delta\tau$ equal for all municipalities. It will most likely be the case that these vary slightly from region to region.
- The number of imported cases in each municipality is defined by a crossing parameter that couples all municipalities. Our model thus does take into account the geography. A simple extension of our model would make the cross parameter $c$ depend on the distance between municipalities. This will likely change the value of $c$, but only slightly affect $R$ and $k$.
- We assume that $R(t)$ varies slowly. Interventions can in principle make sudden changes to $R(t)$.

- To obtain Eq. (9) we assumed independence of the random variables. As mentioned in the main text, this will cause our distribution of $\Delta\tau$ to be too wide. We tested the effect of this by considering narrower distributions.
- We assume that municipalities are uncorrelated except for their testing frequency. After correcting for testing we, nonetheless, measure a correlation coefficient of $\sim 0.01$ as shown in Fig. 1.
